# Single-session intervention with and without video support to prevent the worsening of emotional distress among healthcare workers during the SARS-CoV-2 pandemic: a randomized clinical trial

**DOI:** 10.1101/2024.09.04.24313087

**Authors:** Giovanni Abrahão Salum, Lucas Spanemberg, Marianna de Abreu Costa, André Rafael Simioni, Natan Pereira Gosmann, Lívia Hartmann de Souza, Pim Cuijpers, Daniel Samuel Pine, André Russowsky Brunoni, Natan Katz, Roberto Nunes Umpierre, Christian Haag Kristensen, Gisele Gus Manfro, Marcelo Pio de Almeida Fleck, Carolina Blaya Dreher

**Affiliations:** Hospital de Clínicas de Porto Alegre, Universidade Federal do Rio Grande do Sul - Porto Alegre, RS - Brazil; Child Mind Institute - New York, NY - United States; Pontifícia Universidade Católica do Rio Grande do Sul, Porto Alegre, RS - Brazil; Department of Clinical, Neuro and Developmental Psychology, Amsterdam Public Health Research Institute, Vrije Universiteit Amsterdam, Amsterdam, Netherlands; Emotion and Development Branch, Section on Development and Affective Neuroscience, National Institute of Mental Health, Bethesda, USA; Departamento de Psiquiatria da Faculdade de Medicina da Universidade de São Paulo, São Paulo, Brazil; Instituto de Psiquiatria do Hospital das Clínicas da Faculdade de Medicina da USP, São Paulo, Brazil; Projeto Telessaúde RS, Universidade Federal do Rio Grande do Sul, Porto Alegre, Brazil

## Abstract

**BACKGROUND:** The SARS-CoV-2 pandemic was a major stressful event that significantly affected healthcare providers. This study aimed to compare the efficacy of a single-session intervention (SSI) with and without weekly personalized pre-recorded videos to prevent the worsening of emotional distress.

**METHODS:** Nationwide randomized clinical trial conducted in Brazil from May 19, 2020 to December 31, 2021. We included healthcare professionals with levels of anxiety, depression, or irritability below a T-score of 70, defined by the Patient-Reported Outcomes Measurement Information System. The exclusion criterion was suicide risk. Participants were randomized to SSI or SSI plus weekly personalized pre-recorded videos for four weeks (SSI-ET). The primary outcome was the proportion of participants with a significant escalation of symptoms (i.e., T-score above 70). The trial was registered with ClinicalTrials.gov (NCT04632082).

**FINDINGS:** Of the 3328 volunteers assessed for eligibility, 1112 participants were allocated to either SSI (n=549) or SSI-ET (n=563). The cumulative proportion of incident cases at six months was 17.5% (95% CI 13.1% to 21.6%) for SSI and 15% (95% CI 10.9% to 18.8%) for SSI-ET, with no difference between groups (HR=0.81, 95% CI 0.83 to 1.79). Both groups presented significant symptom reduction in the one-month assessment and maintained over three and six-month follow-ups (Cohen’s d range = 0.68 - 1.08).

**CONCLUSIONS:** We found no benefit of adding asynchronous videos to SSIs; however, we found strong effects for the synchronous component for preventing symptom worsening. This component emphasizes core elements of mental health support, such as active listening, validation, and empathic care. Due to its simplicity and scalability, this intervention shows promise as a public health measure to be widely implemented.

**FUNDING:** Ministry of Health of Brazil, Coordenação de Aperfeiçoamento de Pessoal de Nível Superior, Conselho Nacional de Desenvolvimento Científico e Tecnológico, and Fundo de Incentivo à Pesquisa/Hospital de Clínicas de Porto Alegre.

## INTRODUCTION

The SARS-CoV-2 pandemic has profoundly impacted the mental health of healthcare workers,^1^ underscoring the need for preventive interventions against the worsening of emotional distress. Brazil is among the countries that have been severely affected by the COVID-19 pandemic, with a staggering 38.7 million cases and a mortality rate of 338 deaths per 100,000 inhabitants during the pandemic. The total number of deaths from COVID-19 exceeds 711,000, making Brazil the country with the second-highest number of deaths from COVID-19 worldwide.^2^ In particular, frontline healthcare workers had faced higher risks of adverse mental health outcomes due to increased infection rates, workload, and the psychological strain of patient care.^1^ Therefore, early preventive strategies have been crucial for mitigating the immediate effects of stress and preventing symptom escalation to more severe mental health conditions.^3,4^

During crisis situations, individuals often exhibit symptoms of depression, post-traumatic stress, and anxiety; however, most of them neither develop psychiatric syndromes nor require long-term treatment.^5^ Additionally, psychological debriefing, once a common intervention, has been critically re-evaluated and recent guidelines now advise against offering this technique to trauma-exposed individuals.^6,7^

In this context, brief interventions like ‘Psychological First Aid’ have been widely recognized as the primary choice for initial care, despite limited evidence supporting their effectiveness.^8^ However, the potential benefits of integrating such techniques with routine outcome assessments and risk stratification, a foundation for enhanced psychoeducation, remain largely unexplored. Consequently, the effectiveness of personalized psychoeducation in augmenting simple interventions for individuals in high-exposure crisis scenarios is poorly understood. Also, despite the increasing evidence supporting the efficacy of single-session psychological interventions for the treatment of various conditions, such as depression and anxiety,^9^ the efficacy of these interventions for the prevention of mental disorders remains unassessed.

This study aimed to address these gaps by examining the impact of adding personalized psychoeducational components to a single synchronous session aimed at preventing the worsening of emotional distress among healthcare workers during the SARS-CoV-2 pandemic. Through a nationwide two-arm randomized clinical trial in Brazil, this study compared a single-session intervention (SSI) with an enhanced version incorporating weekly personalized pre-recorded videos for four weeks (single-session intervention enhanced telepsychoeducation, SSI-ET). We hypothesized that the enhanced intervention would significantly reduce the incidence of symptom worsening at six months, based on the premise that a combination of synchronous and asynchronous psychoeducational elements could more effectively manage emotional distress in healthcare workers.

## METHODS

### Study design

The TelePSI trial was a nationwide, investigator-initiated, unicenter, randomized, unblinded, pragmatic controlled trial conducted in Brazil. This trial evaluated the effectiveness of two remote interventions: SSI and SSI-ET. The trial was designed and supervised by a steering committee of members from academia and international experts in emotional disorders (DSP) and psychotherapy for emotional disorders (PC).

The National Research Ethics Commission (CONEP) approved the trial protocol. This article adhered to the ethical principles outlined in the Declaration of Helsinki and the guidelines established in the approved protocol and is reported as recommended by the CONSORT 2010 guideline (appendix pp 2-3). This trial received funding from the Ministry of Health of Brazil through the decentralized execution term No. 16/2020.

### Participants

Participants were recruited nationwide via helplines, e-mails, social media, and traditional media (e.g., national and local newspapers and TV commercials). All assessments and study procedures were conducted online and all participants provided oral informed consent.

The inclusion criteria were the following: (1) professionals or interns from the health service seeking help for their emotional distress in one of our channels; (2) T-score below 70 in any of the Patient-Reported Outcomes Measurement Information System (PROMIS) scales of anxiety, depression or anger.^10^ Importantly, in the protocol defined *a* priori, a z-score on the PROMIS score below 1.5 was included as an inclusion criterion; nevertheless, our recruitment soon revealed that a substantial proportion of participants would have been excluded using this criterion. Thus, this criterion was not used during participants enrollment.

The exclusion criterion was suicidal ideation assessed using one question from the Patient Health Questionnaire (PHQ-9).^11^ Individuals who rated one, two, or three on this question were immediately referred to a psychiatrist to ensure participants’ safety.^12^ There was no restriction for inclusion regarding self-reported sex at birth (i.e., male or female).

### Randomization and masking

The participants were randomly allocated in a 1:1 ratio to the SSI or SSI-ET group. Randomization occurred at the intervention and therapist levels immediately after the participants completed the online questionnaires and was centrally conducted by independent research assistants. An automated computer-generated random number table was used to perform randomization. It automatically assigned participants to one of the two arms based on their trial ID numbers. Due to the nature of the interventions, both participants and therapists were aware of the assigned intervention, but participants were unaware of which intervention was of interest to the researchers.

### Procedures

SSI or SSI-ET sessions started no later than one week after randomization. All sessions were conducted online using Google Meet. At the end of the treatment (week four), we sent online self-report questionnaires to participants via *WhatsApp*. Follow-up assessments were conducted three and six months after treatment. Participants received detailed reports of their answers via email. In cases in which respondents did not respond to the questionnaire links, the research team sent up to six reminders to encourage the completion of the assessments.

### Single-session Intervention

The SSI protocol consisted of a one-session intervention with a trained psychologist or psychiatrist. Therapists were instructed to listen carefully and empathetically to the participants and to create a welcoming, gentle, and non-judgmental environment in which participants felt comfortable and confident in expressing their thoughts, concerns, and emotions. The general principle was to assist participants in stressful situations, encouraging them to develop autonomy, self-efficacy, and problem-solving without harming, strengthening their support network, and developing the ability to feel safe, not alone, calm, and hopeful, as well as to ensure social, physical and emotional support. During the session the therapist first reviewed participants’ symptom levels using scores obtained from the PROMIS Anxiety, Depression, and Anger scales. Soon after, the participants’ responses regarding protective behaviors (e.g., meditating, reading books, and eating healthily) and risky behaviors (e.g., self-medication, excessive alcohol consumption, and problematic use of social media) were reviewed. The PROMIS scale scores and the survey of risk/protective behaviors were shared in the form of visually appealing graphics during the online session. The therapist and patient then negotiated a plan to address mitigating negative coping mechanisms and increase the number of positive coping mechanisms.

### Single-session Intervention Enhanced Telepsychoeducation

The SSI-ET includes all components of SSI, plus two short videos per week that were sent via text message to the participants based on their needs for four weeks. To personalize the treatment, psychoeducation videos were carefully selected by the therapists based on participants’ responses to PROMIS scales, risk/protective behaviors, and their expressed needs during the session. Over the following four weeks, the therapists sent a total of eight videos (two videos per week) to the participants. These videos were selected from a list of 16 that covered the following topics: contagion fear, typical vs. excessive anxiety, typical vs. excessive depression, anger vs. irritability, burnout, acute stress reactions, sleep hygiene, healthy eating habits, exercise, excessive consumption of alcohol and drugs, excessive exposure to news, excessive use of social media, taking care of children, taking care of the elderly, and social support. These videos were carefully developed by the researchers and are freely available on the TelePSI website (https://telepsi.hcpa.edu.br). Additionally, the therapists were available for four weeks to address any messages or queries sent by the participants through a phone chat. The appendix (pp 4-5) presents additional information regarding the session format and video content.

### Outcomes

The primary outcome was the proportion of participants with a T-score ≥ 70 on the Patient-Reported Outcomes Measurement Information System (PROMIS) scales of anxiety, depression, and/or irritability at six months.

The secondary outcomes were the proportion of participants with a T score ≥ 60 on any of the emotional distress scales at one, three, and six months; and mean score changes between groups in anxiety, depression, and irritability. Other secondary outcomes were life satisfaction, measured using the Satisfaction with Life Scale, which captures individuals’ appraisal of their overall well-being and contentment with life and reports its results with higher scores indicating higher satisfaction with life, and service satisfaction assessed through the net-promoter score (NPS), calculated by asking participants a single question regarding the likelihood of recommending each therapy to someone on a scale from 0 to 10. The respondents were classified as promoters (9-10), passives (7-8), or detractors (0-6). The NPS was derived by subtracting the percentage of detractors from promoters by coding detractors as −1, passives as 0, and promoters as 1. We also asked participants to rate their overall satisfaction using a single question ranging from 1 (’Extremely dissatisfied’) to 7 (’Extremely satisfied’). Measures of anxiety, depression, irritability (i.e., the primary outcomes), and life satisfaction were measured at months one, three, and six, and service satisfaction was evaluated at month one.

Participants’ safety was ensured by tracking suicide attempts, psychiatric hospitalization, and a short questionnaire developed by the research team to track the side effects of psychosocial interventions with the following items: “*My mental health is worse because of therapy*”, “*I developed a dependency on therapy to get on with my life*”, “*Therapy made me feel guilty about my problems*”, “*Therapy reactivated bad thoughts that worsened my mental health*”, “*I got involved in unnecessary conflicts with other people because of therapy*”, “*Therapy led me to think there is no solution for my problems*”, “*Therapy made me feel unable to face my problems*”, and “*Overall, therapy has done me more harm than good*”. Participants were required to respond to each statement using a 5-point Likert scale, which measured the extent to which they agreed or disagreed with the statement. For analysis, the responses were subsequently categorized into two groups: those who indicated the presence of undesirable side-effects (”Completely Agree” or “Agree”) and those who did not (”Completely Disagree”, “Disagree”, or “Neither agree nor disagree”).

The therapists’ fidelity to each protocol was assessed by a supervisor who reviewed a series of recorded sessions that were randomly selected from each therapist. The supervisor used a scale developed by our team to evaluate the quality and fidelity of each session. In addition, the supervisor met the therapists in group supervision weekly to ensure protocol fidelity. The assessment of the session quality involved seven distinct criteria. Among these, seven skills are overarching and common to various interventions: (1) establishing a therapeutic alliance with the patient; (2) structuring and organizing the session; (3) compassion, empathy, genuineness, and warmth; (4) effective use of non-specific therapeutic skills such as clarification, reflection, and validation; (5) confidence and self-assurance; (6) reassurance; and (7) proficiency in instilling hope. For therapists practicing SSI-ET, assessment focused exclusively on general skills. Each item was rated on a scale of 1 (low) to 5 (high), with higher scores indicating greater session fidelity.

### Choice of the primary outcome

The primary outcome measures were PROMIS anxiety, depression, and anger scales. PROMIS is a self-rated assessment tool developed by the National Institutes of Health (NIH) to assess patients’ perspectives on their health status and overall quality of life. These scales, covering emotional states over the previous seven days, comprised eight items for anxiety, eight items for depression, and five items for anger. Responses are recorded on a 5-point Likert scale (1=’Never’, 2=’Rarely’, 3=’Sometimes’, 4=’Often’, 5=’Always’), with higher scores indicating increased emotional distress. All PROMIS scales have high internal consistency and reliability.^10^ We used PROMIS T-scores with an average of 50 and a standard deviation of 10. Therefore, a T-score of 70 (i.e., our inclusion criterion) represents 2 standard deviations above the sample mean. This measure was chosen as the primary outcome because it is designed to be patient-friendly and uses fewer items than most scales designed to evaluate the same symptom domains, thereby decreasing the respondent burden. Additionally, this instrument is freely available and has been translated into several languages.

### Statistical analysis

The study was designed to have 90% power to detect a 5% difference between groups in the primary outcome (alpha 0.05). Estimating a 20% loss to follow-up required a total of 1100 participants. Analysis was performed according to intention-to-treat criteria (ITT).

The cumulative proportion of incident cases at 6 months was assessed using survival analysis with all available data. Differences between groups and hazard ratios were estimated using Cox proportional hazards. Continuous outcomes were analyzed using linear mixed models with all available data from baseline, one, three, and six-month follow-up. Comparisons of NPS, overall treatment satisfaction, and adverse events between groups were tested using a Multiple Imputation Welch Two Sample t-test and pooled results from logistic regressions accounting for missing data. All analyses were performed using R. Survival analysis and linear mixed models, equivalence tests, and parameters were conducted or estimated using *survival*, *lme4*, *mice*, *MKmisc*, and *Effects* packages.^13–15^ Cohen’s *d* was used as a measure of standard effect size for significant associations, with values of 0.2, 0.5, and 0.8 interpreted as small, medium, and large, respectively.^16^

In order to formally test for the absence of minimal clinically relevant differences between treatment arms, we have also included, a posteriori, an equivalence test (two one-sided tests equivalence testing, TOST) between interventions considering −7% as the smallest effect size of interest, as defined a priori for the statistical power analysis.^17^ This represents approximately half of the differences found for the other prevention trials for the same time period.^18^

Due to the crisis context of the SARS-CoV-2 pandemic, this trial was prospectively registered at ClinicalTrials.gov (NCT04632082) on November 11, 2020, during the recruitment process; nevertheless, less than 6% of participants had already been included by the time of registration, no preliminary statistical analyses were performed, and only minor changes in the previously defined methodological procedures were implemented during the entire trial. Rather than excluding participants based on a clinical assessment of suicide, we excluded them based on affirmative responses to the PHQ-9 suicide item. This decision stemmed from the concern that randomizing participants after the psychiatric assessment might introduce bias in a trial designed to compare distinct interventions against a single session. Also, a z-score on the PROMIS score below 1.5 was initially defined as an inclusion criterion; however, our recruitment process soon revealed that a substantial proportion of participants would have been excluded using this criterion and this criterion was not implemented.

## RESULTS

Of the 3328 individuals who sought to participate in this project from May 19, 2020, to December 31, 2021, 3086 consented to participate in the study, and 490 were excluded and subsequently referred to a psychiatrist due to concerns about suicide risk. We enrolled 1112 participants who met the inclusion criteria and were subsequently allocated to SSI (n=549) and SSI-ET (n=563) (Figure 1). The majority of the sample consisted of women (86.4%), and the mean age of the sample was 37.2 (SD=9.46). Each group is further described in Table 1.

**Figure 1.**
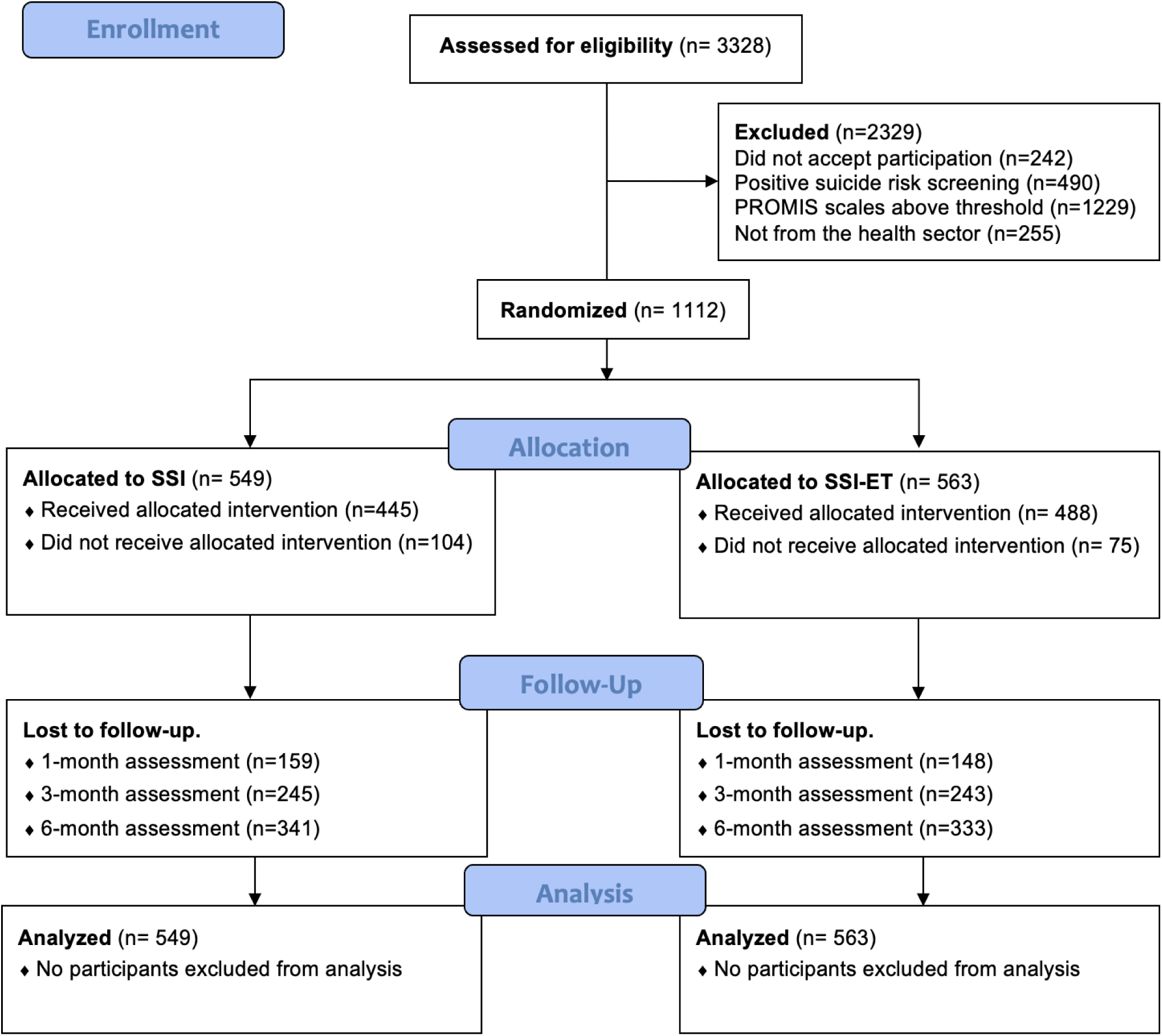
CONSORT Flow Diagram

**Table 1.** Sample description among groups.

| .. | **SSI**<br>(n=549) | **SSI-ET**<br>(n=563) |
| --- | --- | --- |
| **Sex at birth (female)** | 475 (86.7%) | 485 (86.1%) |
| **PROMIS Anxiety** | .. | .. |
| None to slight | 27 (4.9%) | 15 (2.7%) |
| Mild | 64 (11.7%) | 71 (12.6%) |
| Moderate | 458 (83.4%) | 477 (84.7%) |
| **PROMIS Depression** | .. | .. |
| None to slight | 143 (26%) | 140 (24.9%) |
| Mild | 193 (35.2%) | 193 (34.3%) |
| Moderate | 213 (38.8%) | 230 (40.9%) |
| **PROMIS Irritability** | .. | .. |
| None to slight | 214 (39%) | 204 (36.2%) |
| Mild | 105 (19.1%) | 121 (21.5%) |
| Moderate | 230 (41.9%) | 238 (42.3%) |
| **Age (years) - Mean (SD)** | 37.5 (9.06) | 36.9 (9.84) |
| **PROMIS Anxiety (T-score) - Mean (SD)** | 64.2 (4.80) | 64.5 (4.45) |
| **PROMIS Depression (T-score) - Mean (SD)** | 57.9 (5.68) | 58.3 (5.84) |
| **PROMIS Irritability (T-score) - Mean (SD)** | 57.4 (7.77) | 57.4 (7.71) |
| **Satisfaction with Life (sum) - Mean (SD)** | 20.4 (5.91) | 19.9 (5.68) |

PROMIS, Patient-Reported Outcomes Measurement Information System; SSI, single-session intervention; SSI-ET, single-session intervention - enhanced psychoeducation; SD, standard deviation.

### Primary outcome

The estimated cumulative proportion of incident severe cases at 1 month was 6.9% (95% CI 9.4% to 4.4%) in the SSI group and 4.8% CI (95%CI 6.9% to 2.7%) in the SSI-ET group, at 3-months it was 12.4% (95% CI 15.8% to 8.9%) in the SSI group and 9.9% (95% CI 12.9% to 6.8%) in the SSI-ET group, and at 6-months it was 17.5% (95% CI 13.1% to 21.6%) in the SSI group and 15% (95% CI 10.9% to 18.8%) in the SSI-ET group with no difference between groups (HR=0.81, 95% CI 0.83 to 1.79, p=0.291) (figure 2).

**Figure 2.**
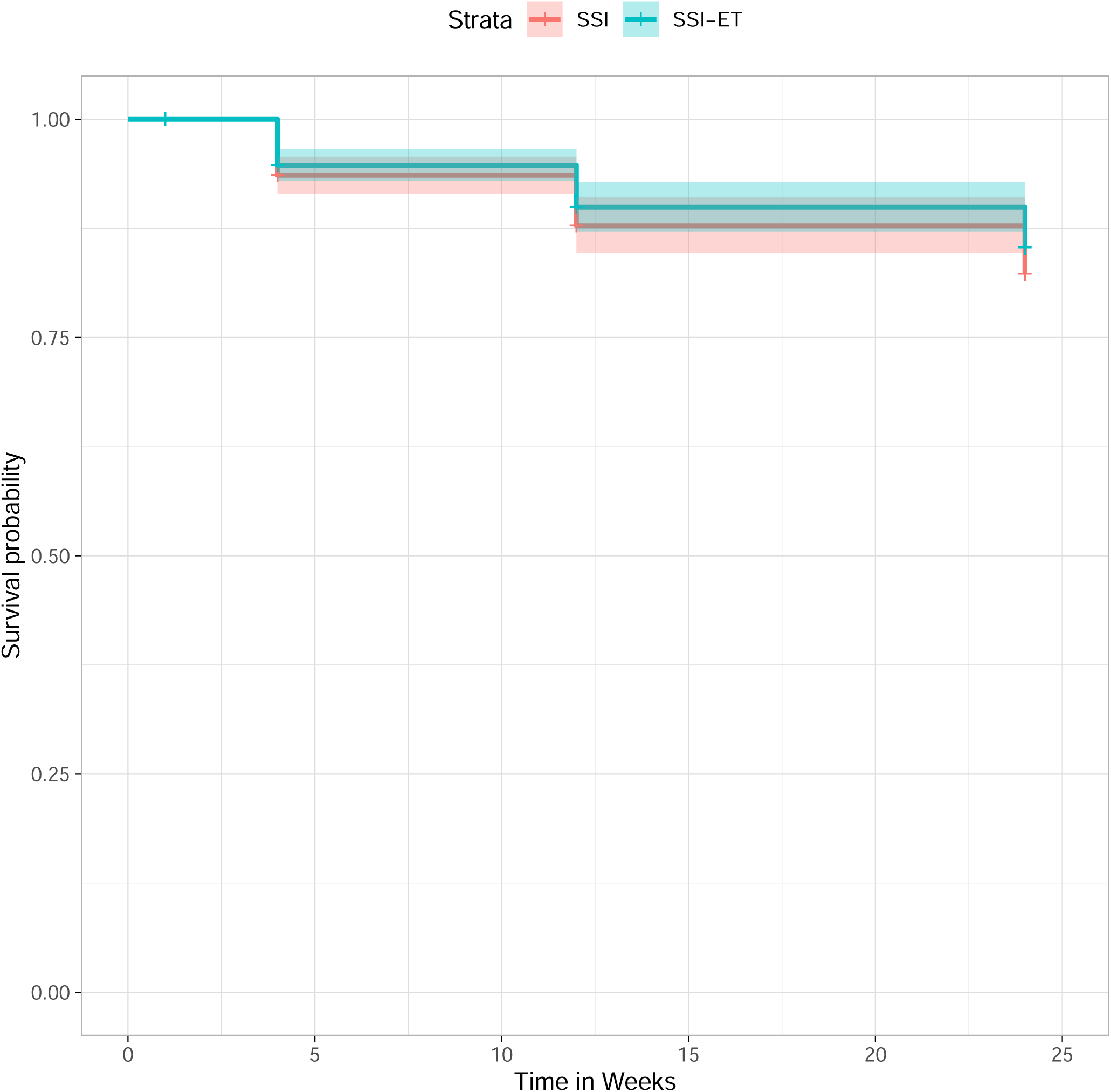
Survival analysis. Legend: SSI, Single-Session Intervention; SSI-ET, SSI with Enhanced Psychoeducation.

### Secondary outcomes

#### Incidence of subthreshold levels of emotional distress (T-scores above 60)

This analysis was performed using the sample that did not have a score of 60 or above at baseline (n=143). The estimated cumulative proportion of incident severe cases at 1 month was 20.8% (95% CI 31.5% to 8.5%) in the SSI group and 19.4% (95% CI 17% to 21.2%) in the SSI-ET group, at 3-months it was 25.2% (95% CI 36.6% to 11.8%) in the SSI group and 25.7% (95% CI 37.3% to 11.9%) in the SSI-ET group, and at 6-months it was 32.7% (95% CI 45.1% to 17.5) in the SSI group and 28.9% (95% CI 41.3% to 13.9%) in the SSI-ET group, with no difference between groups (HR=0.81, 95% CI 0.58 to 2.53, p=0.587).

#### Relevant time effects

Linear mixed models, utilizing all available data, showed prominent time effects for all outcomes, indicating significant differences from baseline to the one-, three-, and six-month follow-up assessments (table 2). The mean changes were significant in each treatment arm. Standardized symptom reduction for anxiety symptoms from baseline to one-month was Cohen’s d=0.68 (p<0.001) for SSI and Cohen’s d=0.77 (p<0.001) for SSI-ET. Standardized symptom reduction for depressive symptoms from baseline to one-month was Cohen’s d=0.86 (p<0.001) for SSI and Cohen’s d=1.06 (p<0.001) for SSI-ET. Finally, standardized symptom reduction for irritability symptoms from baseline to one-month was Cohen’s d=0.68 (p<0.001) for SSI and Cohen’s d=0.89 (p<0.001) for SSI-ET. All differences were maintained at the 3- and 6-month follow-ups with similar magnitudes (figure 3).

**Figure 3.**
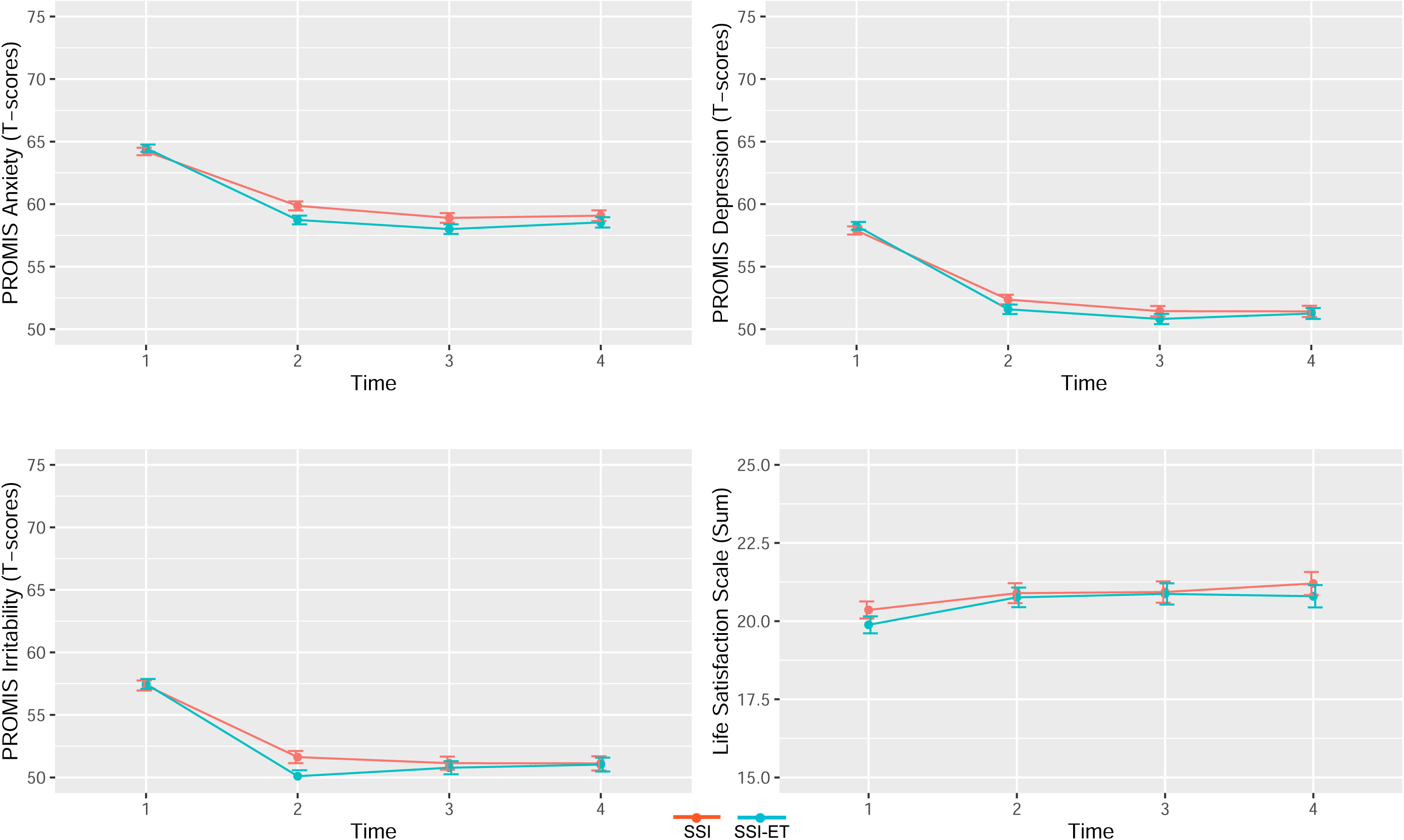
Estimated scores on all secondary outcomes as a function of each timepoint (baseline, one-month, three-months, and six-months follow-up). Legend: SSI, Single Session Intervention; SSI-ET, SSI with Enhanced Psychoeducation.

**Table 2.**
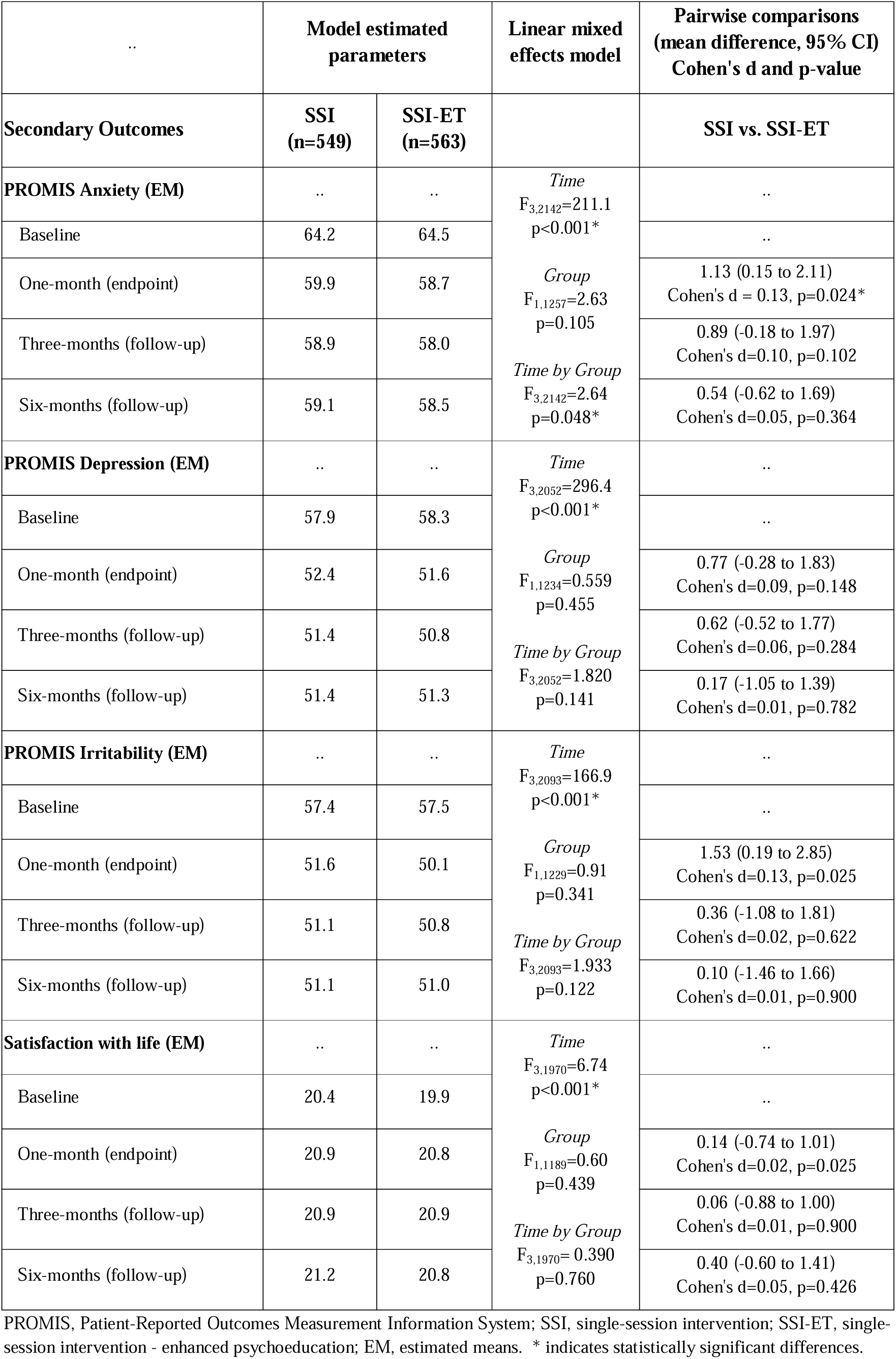
Linear Mixed Models hypothesis testing, parameter estimates and pairwise comparisons.

#### Time-by-group interactions

We found significant time-by-group interaction for the PROMIS anxiety scores over time (Table 2), meaning that at 1-month SSI was significantly worse than SSI-ET in reducing PROMIS anxiety scores (mean difference=1.12, 95% CI 0.14 to 2.11, p=0.024; Cohen’s d=0.14). The difference was no longer significant at the 3-month or 6-month follow-ups (Cohen’s d range 0.05-0.09, p-values>0.05). No time-by-group interactions were observed for irritability or depressive symptoms.

#### Net Promoter Scores and overall satisfaction

There were no significant differences in the probability of participants recommending the treatment between treatment arms. The estimated NPS for SSI was 53% and that for SSI-ET was 54.6% (estimated mean difference=2.6%, t=0.339, df=27, p-value=0.737). There were also no differences between the overall treatment satisfaction with scores of 5.6 for SSI and 5.7 for SSI-ET (estimated mean difference=0.12, t=1.125, df=103, p-value=0.213).

#### Therapists’ fidelity

The supervisor observed ten psychoeducation sessions without videos and nine sessions with videos. The average fidelity score for therapists leading the SSI was 4.94 (SD 0.1), while for therapists facilitating the SSI-ET, the mean score was 4.78 (SD 0.32), with no difference between groups (p>0.05).

#### Attrition

The flow diagram in Figure 1 illustrates the number of individuals assessed at one-, three-, and six-month follow-up, demonstrating considerable attrition rates. There was a significant between-group differences in the proportion of participants that did not receive the allocation intervention between groups (18.9% in SSI vs. 13.3% in SSI-ET, □2=6.16, df=1, p-value=0.013). A sensitivity analysis using modified intention to treat (excluding those who did not receive allocated intervention) revealed similar results for all outcomes (primary outcome: HR=0.807, p=0.269). There were no significant differences between losses in follow-up between groups (□2=0.958, df=3, p-value=0.8114)

#### Post-hoc equivalence tests

Equivalence tests indicating the observed difference between each pair of interventions were within the equivalence bounds of −7% and 7% for the primary outcome: SSI vs. SSI-ET (Z=- −1.81, p=0.035).

### Safety assessment

A total of three patients attempted suicide in the 1-month follow up (SSI: 2/349, 0.6% vs. SSI-ET:1/367, 0.3%, □^2^= 0.39, p=0.534), one at the 3-months follow up (SSI:1/288, 0.3% vs. SSI-ET: 0/292, 0%, □^2^= 1.02, p=0.314) and three at 6-months follow-up (SSI: 1/235, 0.4% vs. SSI-ET: 2/250, 0.8%, □^2^= 0.28, p=0.599). A total of nine patients were hospitalized for mental health reasons in the 1-month follow-up (SSI: 4/349, 1.1% vs. SSI-ET:5/367, 1.4%, □^2^= 0.07, p=0.795), seven at the 3-months follow-up (SSI:2/288, 0.7% vs. SSI-ET: 5/292, 1.7%, □^2^= 1.26, p=0.262) and four at 6-months follow-up (SSI: 1/235, 0.4% vs. SSI-ET: 3/250, 1.2%, □^2^= 0.89, p=0.346). No differences were observed between the SSI and SSI-ET groups in terms of undesirable side effects related to the intervention, as shown in appendix p 5.

## DISCUSSION

In this nationwide two-arm randomized clinical trial conducted in Brazil, we investigated the efficacy of single-session interventions (SSIs) over the SSIs enhanced with personalized psychoeducational videos (SSI-ET) to prevent the worsening of emotional distress among healthcare workers during the SARS-CoV-2 pandemic and found no significant advantage of the video-enhanced SSIs on preventing emotional distress exacerbation. These results reinforce the importance of the synchronous components of SSIs for preventing symptom worsening in healthcare workers. Both groups showed a reduction in symptoms from baseline to endpoint with substantial effect sizes for this population and this improvement was sustained over the three- and six-months follow-ups. This finding is critical as it substantiates the effectiveness of SSIs as a standalone intervention in a high-stress cohort, adding to the evidence of the value of mental health interventions during crises.

The intervention, grounded in Psychological First Aid principles and augmented with measurement-based care and risk stratification components, aimed to provide a more holistic approach to crisis mental health support. Despite the wide application of Psychological First Aid, the empirical evidence of its effectiveness remains limited.^19^ Our study suggests that the core principles of Psychological First Aid are essential and that the additional psychoeducational components with therapist contact do not significantly enhance intervention efficacy. The role of single session interventions in psychological interventions is increasing across various conditions, including depression and anxiety.^9^ Our study extends this evidence to the preventative domain, demonstrating that SSIs can effectively reduce the symptoms of a population with moderate levels of emotional distress, and not only to those with severe levels of symptoms. While we do not know the baseline incidence levels of depression in this population without active interventions, previous studies have shown an approximate rate of 25-30% in control interventions at 24 weeks follow-up, which is substantially higher than 15-17.5% we found here.^18^ While these studies are not directly comparable, they provide some benchmarks for what would be expected in a six-month period for participants with subthreshold symptoms.

Regarding the undesirable side effects of psychotherapy, the incidence was low for all measures, except for the concern of dependence (above 5%). This concern was raised by the participants even though the therapist had very limited contact. The contact with the therapist on sending the videos did not increase patient satisfaction, which again reinforced the importance of synchronous contact with the therapist. Safety assessment during the follow-up period found that seven participants attempted suicide and 20 participants were hospitalized for mental health reasons. The majority of suicide attempts in our study occurred during the initial month of follow-up and reinforce the need of further assessment for suicidality after therapy.

This study has important limitations. First, we had a significant attrition rate and reliance on virtual environment self-reports, which may have impacted the representativeness and comparability of the findings with the current literature. Despite these challenges, methodological rigor was maintained through statistical approaches that accounted for all available data and used validated patient reported outcomes. The absence of a control group, dictated by ethical considerations, underscores the need for further research exploring the comparative effectiveness of such interventions. In addition, the videos were freely accessible on our website, therefore, we cannot guarantee that contamination did not occur in the SSI arm.

In summary, this study offers important evidence for the efficacy of SSIs in preventing emotional distress among healthcare workers during a pandemic, emphasizing the synchronous component of the intervention. These findings have significant implications for the design and implementation of mental health interventions in crises, advocating for the adoption of SSIs due to their simplicity, effectiveness, and ease of scalability. Future research should continue to explore the potential of brief, transdiagnostic interventions in diverse settings, particularly focusing on scalable solutions for mental health support during crises.

## Contributors

GAS and CBD conceived and designed the study. GAS and CBD obtained funding for the trial. GAS, MAC, LS, ARS, NPG, and LHS provided administrative, technical, or material support. GAS developed the statistical analysis plan. GAS, AS, and NPG conducted the statistical analysis. GAS, PC, DSP, NK, RNU, ARB, CHK, MPAF, GGM, and CBD supervised the conduction of the trial and production of this Article. All authors drafted and critically revised the content of this article. All authors have read and approved the final manuscript, confirm that they had full access to the data, and accept responsibility to submit for publication.

## Data sharing

Individual participant data that underlie the results reported in this article, after de-identification (text, tables, figures, and appendices) and the study protocol is available immediately following publication with no end date (appendix pp 6-18). Researchers who provide a methodologically sound proposal may gain access to the database to achieve aims in the approved proposal. Proposals should be directed to and, to gain access, data requestors will need to sign a data access agreement. After 36 months, data will be available for 5 years at our university’s data warehouse (https://lume.ufrgs.br), but without investigator support other than deposited metadata.

## Acknowledgment

This study was financed in part by the Ministry of Health of Brazil through, the decentralized execution term No. 16/2020, Coordenação de Aperfeiçoamento de Pessoal de Nível Superior (CAPES), Finance Code 001, Conselho Nacional de Desenvolvimento Científico e Tecnológico (CNPq - 001), Brazilian federal government agencies, and Fundo de Incentivo à Pesquisa/Hospital de Clínicas de Porto Alegre (FIPE/HCPA - 001) – Brazil. The funders of the study had no role in the design or conduct of the study; collection, management, analysis or interpretation of the data; preparation, review or approval of the manuscript; or decision to submit the manuscript for publication. The views expressed are those of the authors and not necessarily those of the Ministry of Health of Brazil, CAPES, CNPq, and FIPE/HCPA. This project was only possible due to the effort and collaboration of many people and institutions; therefore, we would like to thank our collaborators from Hospital de Clínicas de Porto Alegre, Pontifícia Universidade Católica do Rio Grande do Sul, Universidade Federal de Ciências da Saúde de Porto Alegre, Universidade de São Paulo, Universidade Federal de São Paulo, Pan American Health Organization/WHO, Sociedade Brasileira de Psicologia, Associação de Psiquiatria do Rio Grande do Sul, Universidade Federal do Paraná, Universidade Estadual de Campinas, Centro de Estudos em Psiquiatria Integrada, Liga de Psiquiatria e Saúde Mental da UFCSPA/UFRGS, The Funn Club, Yoho Arsenal, and Agência LVL Funding.

