## appendix for "Single-session intervention with and without video support to prevent the worsening of emotional distress among healthcare workers during the SARS-CoV-2 pandemic: a randomized clinical trial"

**Supplementary Appendix**

**Contents**

**Appendix 1: CONSORT checklist (pg. 2)**

**Appendix 2: Protocol of the single session intervention-enhanced telepsychoeducation group (SSI-ET) (pg. 4)**

**Appendix 3: Therapy-specific adverse effects (pg. 5)**

**Appendix 4: Study protocol (pg. 6)**

**Appendix 1: CONSORT checklist**

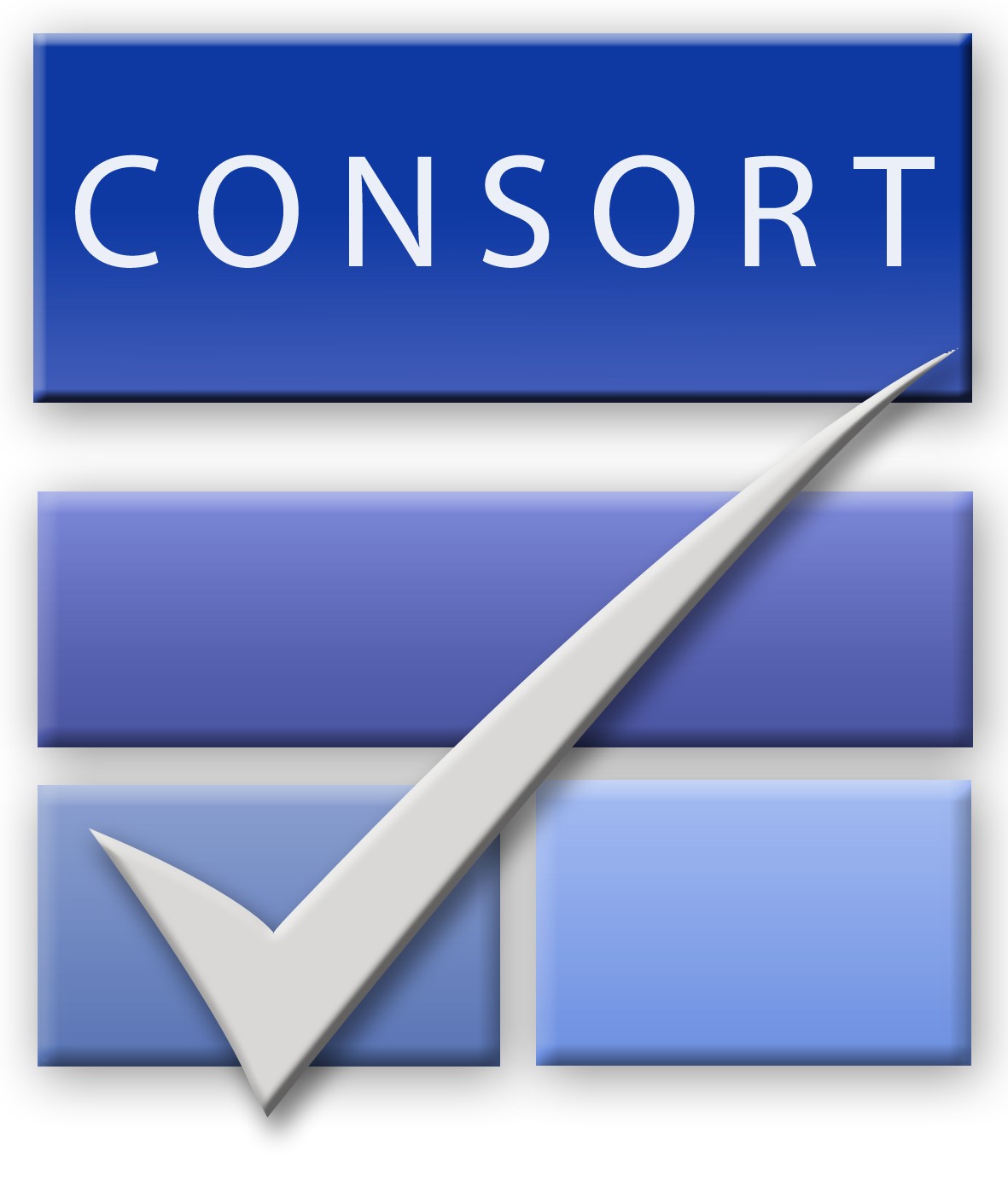

**CONSORT 2010 checklist of information to include when reporting a randomised trial***

| **Section/Topic** | **Item No** | **Checklist item** | **Reported on page No** |
| --- | --- | --- | --- |
| **Title and abstract** | | | |
| ™ | 1a | Identification as a randomised trial in the title | 1 |
|  | 1b | Structured summary of trial design, methods, results, and conclusions (for specific guidance see CONSORT for abstracts) | 4 |
| **Introduction** | | | |
| Background and objectives | 2a | Scientific background and explanation of rationale | 5 |
|  | 2b | Specific objectives or hypotheses | 5 |
| **Methods** | | | |
| Trial design | 3a | Description of trial design (such as parallel, factorial) including allocation ratio | 6 |
|  | 3b | Important changes to methods after trial commencement (such as eligibility criteria), with reasons | 11 |
| Participants | 4a | Eligibility criteria for participants | 6 |
|  | 4b | Settings and locations where the data were collected | 6 |
| Interventions | 5 | The interventions for each group with sufficient details to allow replication, including how and when they were actually administered | 7-8; appendix |
| Outcomes | 6a | Completely defined pre-specified primary and secondary outcome measures, including how and when they were assessed | 8-9 |
|  | 6b | Any changes to trial outcomes after the trial commenced, with reasons | 8-9 |
| Sample size | 7a | How sample size was determined | 10 |
|  | 7b | When applicable, explanation of any interim analyses and stopping guidelines | 10 |
| Randomisation: |  |  | 7 |
| Sequence generation | 8a | Method used to generate the random allocation sequence |  |
|  | 8b | Type of randomisation; details of any restriction (such as blocking and block size) | 7 |
| Allocation concealment mechanism | 9 | Mechanism used to implement the random allocation sequence (such as sequentially numbered containers), describing any steps taken to conceal the sequence until interventions were assigned | 7 |
| Implementation | 10 | Who generated the random allocation sequence, who enrolled participants, and who assigned participants to interventions | 7 |
| Blinding | 11a | If done, who was blinded after assignment to interventions (for example, participants, care providers, those assessing outcomes) and how | 7 |
|  | 11b | If relevant, description of the similarity of interventions | 7-8; appendix |
| Statistical methods | 12a | Statistical methods used to compare groups for primary and secondary outcomes | 10 |
|  | 12b | Methods for additional analyses, such as subgroup analyses and adjusted analyses | 10 |
| **Results** | | | |
| Participant flow (a diagram is strongly recommended) | 13a | For each group, the numbers of participants who were randomly assigned, received intended treatment, and were analysed for the primary outcome | 11; Fig. 1 |
|  | 13b | For each group, losses and exclusions after randomisation, together with reasons | Fig. 1 |
| Recruitment | 14a | Dates defining the periods of recruitment and follow-up | 11 |
|  | 14b | Why the trial ended or was stopped | 10 |
| Baseline data | 15 | A table showing baseline demographic and clinical characteristics for each group | Table 1 |
| Numbers analysed | 16 | For each group, number of participants (denominator) included in each analysis and whether the analysis was by original assigned groups | Table 1 |
| Outcomes and estimation | 17a | For each primary and secondary outcome, results for each group, and the estimated effect size and its precision (such as 95% confidence interval) | 12-13; Tables 2-3 |
|  | 17b | For binary outcomes, presentation of both absolute and relative effect sizes is recommended | 12-13; Tables 2-3 |
| Ancillary analyses | 18 | Results of any other analyses performed, including subgroup analyses and adjusted analyses, distinguishing pre-specified from exploratory | 12-13 |
| Harms | 19 | All important harms or unintended effects in each group (for specific guidance see CONSORT for harms) | 13 |
| **Discussion** | | | |
| Limitations | 20 | Trial limitations, addressing sources of potential bias, imprecision, and, if relevant, multiplicity of analyses | 15 |
| Generalisability | 21 | Generalisability (external validity, applicability) of the trial findings | 14-15 |
| Interpretation | 22 | Interpretation consistent with results, balancing benefits and harms, and considering other relevant evidence | 14-15 |
| **Other information** | | |  |
| Registration | 23 | Registration number and name of trial registry | 11 |
| Protocol | 24 | Where the full trial protocol can be accessed, if available | 16 |
| Funding | 25 | Sources of funding and other support (such as supply of drugs), role of funders | 16 |

**Appendix 2: Protocol of the single session intervention-enhanced telepsychoeducation group (SSI-ET)**

The enhanced psychoeducation protocol is a one-session intervention based on supportive principles delivered by video call. The general principle is assisting participants in stressful situations using components of psychological first aid. Therapists are oriented to respect participants' safety, dignity, and rights, adapt the intervention to cultural issues, and encourage participants to develop autonomy and self-efficacy. Moreover, the therapists should listen carefully and empathetically to the participants and create a welcoming, gentle, and non-judgmental environment where participants feel comfortable and confident to express their thoughts, concerns, and emotions. Therapists should help participants resolve problems without harming them, strengthen their support network, and develop the ability to feel safe, close to people, calm, and hopeful, as well as ensure social, physical, and emotional support. Improving the coping of the participants, generating greater self-confidence, improving satisfaction, and reducing anxiety, depression, and emotional pain are the main targets of the intervention. The principles of observing, listening, and connecting guide the entire session.

The psychoeducation session has four objectives: (1) offering a safe environment for the participant to express their emotions, and thoughts and share their experiences (10 to 30 minutes); (2) reviewing with the participant the symptomatic assessment which was self-rated by the participant in the project' registration, helping him/her on the areas of greatest concern (5 to 15 minutes); (3) planning lifestyle changes, promoting protective factors and reducing risk factors for mental health; (4) planning the use of psychoeducation videos that will be used over the next four weeks (in the intervention with support videos; ten minutes).

Empathetic listening is the essential component of the entire session, and the main technique adopted in this modality. Listening carefully to the participant's demand, validating emotions and efforts, demonstrating the therapist is attentive, and using verbal and non-verbal components of empathy are the basis of empathetic listening. Other interviewing techniques are encouraged: a. facilitation (refers to the posture, actions, or words that encourage the participant to communicate); b. echo (repetition of participant's words encouraging him/her to express feelings); c. empathic responses; d. validation (the therapist should look for what can be validated in the participant's speech); e. tranquilization; f. summarizing the participant's narrative; g. highlighting transitions in the sessions. h. adapting the language for the participant.

In the intervention with supportive videos, the videos are personalized based on the demands identified in the questionnaire answered by the participant and during the session. The therapists selected eight videos, and sent them to the participant for four weeks (two videos per week), considering the best order and moment to send them (during day-time). Moreover, the therapist was available during this period to answer participants’ messages by phone chat. The following themes comprise each video: 1. SARS-CoV2: How to protect yourself from SARS-CoV2 when you arrive at your home?; 2. Fear of contagion; 3. Normal Anxiety vs. Excessive Anxiety; 4. Sadness vs. Depression; 5. Anger vs. irritability; 6. Burnout: What is Burnout?; 7. Stress and acute reaction to stress; 8. Sleep: Sleep hygiene; 9. Food: Healthy eating and mental health; 10. Exercise: Exercise and mental health; 11. Substances; 12. News: How to protect yourself from excessive news exposure? 13. Social Networks: Excessive use of social networks; 14. Children: How to deal with your children in quarantine times?; 15. Elderly: How to deal with elderly people in quarantine times? 16. Social support: How to stay connected in quarantine times?

**Appendix 3: Therapy-specific adverse effects**

|  | **SSI**  **(n=549)** | **SSI-ET**  **(n=563)** | **OR** | **p-value** |
| --- | --- | --- | --- | --- |
| My mental health is worse because of therapy | 1.38% | 0.74% | 1.87 | 0.549 |
| I developed a dependency on therapy to get on with my life | 5.85% | 8.61% | 0.66 | 0.158 |
| Therapy made me feel guilty about my problems | 1.99% | 3.86% | 0.51 | 0.268 |
| Therapy reactivated bad thoughts that worsened my mental health | 3.5% | 3.2% | 1.09 | 0.847 |
| I got involved in unnecessary conflicts with others because of therapy | 3.12% | 2.46% | 1.27 | 0.577 |
| Therapy led me think there is no solution for my problems | 3% | 3.19% | 0.94 | 0.901 |
| Therapy made me feel unable to face my problems | 2.72% | 1.68% | 1.63 | 0.304 |
| Overall, therapy has done me more harm than good | 2.45% | 1.44% | 1.72 | 0.285 |

SSI, single-session intervention; SSI-ET, single-session intervention - enhanced psychoeducation; OR, Odds ratio.

**Appendix 4: Study protocol**

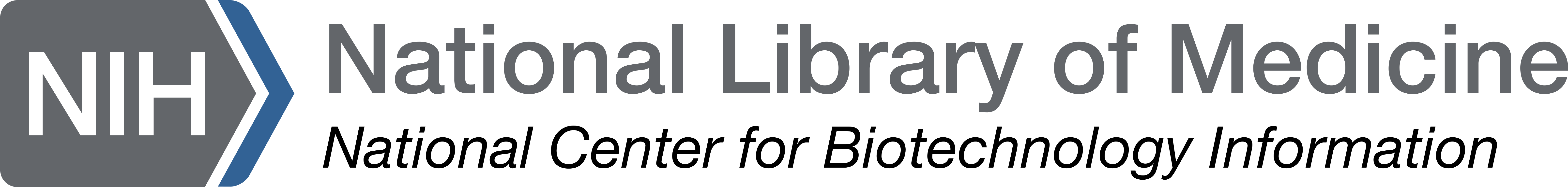
[Go to the classic website](https://classic.clinicaltrials.gov/)

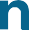

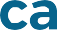

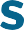

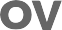

**Record 1 of 1**

**The U.S. government does not review or approve the safety and science of all studies listed on this website.**

Read our full [disclaimer](https://clinicaltrials.gov/about-site/disclaimer) [(https://clinicaltrials.gov/about-site/disclaimer)](https://clinicaltrials.gov/about-site/disclaimer) for details.

UNKNOWN STATUS
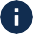

**Psychotherapy Strategies for the Treatment of Professionals and Students From Essential Services With High Levels of Emotional Distress in the Context of COVID-19**

**ClinicalTrials.gov ID**
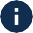
 **NCT04635618**

**Sponsor**
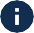
 Hospital de Clinicas de Porto Alegre

**Information provided by**
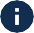
 Giovanni Abrahao Salum Junior, Hospital de Clinicas de Porto Alegre (Responsible Party)

**Last Update Posted**
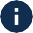
 2020-11-19

**Study Details Tab**

**Study Overview**

**Brief Summary**

The study consists in a pragmatic superiority randomized controlled trial comparing different strategies of psychotherapy for professionals and students from essential services with high levels of emotional distress during the COVID-19 pandemic in Brazil. Therapeutic strategies to be evaluated are Brief Cognitive Behavioral Telepsychotherapy, Brief Interpersonal Telepsychotherapy and Telepsychoeducation, as an active control.

Note: This study was approved by the Ethics and Research Committee of the Hospital de Clínicas de Porto Alegre and is originally registered at Plataforma Brasil, a Brazilian study registration platform

(under CAAE: 30608420.5.0000.5327). Recruitment began in May 28th 2020.

**Detailed Description**

PRAGMATIC TREATMENT TRIAL

TITLE: "A pragmatic superiority randomized controlled trial comparing Brief Cognitive Behavioral Telepsychotherapy, Brief Interpersonal Telepsychotherapy and Telepsychoeducation for the reduction of emotional distress during COVID-19 outbreak in professionals and students from essential services with a high level of emotional distress in Brazil".

IMPORTANCE: COVID-19 outbreak is associated with increased emotional distress (depression, anxiety, and irritability) all over the world. Currently, there are no large randomized trials testing interventions to reduce the burden caused by mental disorders during a pandemic outbreak of these proportions.

OBJECTIVE: To test the effectiveness of two modalities of Brief-Telepsychotherapy (Cognitive Behavioral and Interpersonal) to the reduction of symptoms of emotional distress (anxiety, depression, and irritability) in professionals and students from essential services with a high level of those symptoms in Brazil during the COVID-19 outbreak.

DESIGN, SETTING, AND PARTICIPANTS Thee-arm randomized clinical trial. Participants were recruited in Brazil from the national service of telehealth provided by the ministry of health. Participants included professional and students from essential services suffering from high levels of anxiety, depression, and irritability symptoms during the COVID-19 outbreak. High levels of symptoms were defined by either of the following: (1) T score higher than 70 on the PROMIS Anxiety Scale; (2) T score higher than 70 on the PROMIS Depression Scale; (3) T score higher than 70 on the PROMIS Anger Scale.

INTERVENTIONS: All participants will be randomized 1:1:1: to the Cognitive Behavioral Brief- Telepsychotherapy group (B-CBT, four sessions), Brief Interpersonal Telepsychotherapy (B-IPT, four sessions) or Telepsychoeducation group (a single session psychoeducation group plus weekly personalized pre-recorded videos for 4 weeks).

MAIN OUTCOMES AND MEASURES: The primary outcome will be the proportion of participants with a 50% reduction in T-scores in all the scales that were scored above 70 at baseline at 1-month.

Secondary outcomes (1) the same measure of the primary outcome but measured at 3-month and 6-months follow-up; (2) mean score change in individual scales, quality of life and remission levels (proportion of patients with T-score of 50 or below in all of the four emotional distress subscales);

(3) the same measure of the primary outcome but measured at midpoint (after the second session or 2-weeks); and (4) service satisfaction and net-promoter score at the end of the treatment.

EXPECTED RESULTS: To detect a 15% group difference between each group, an alpha of 0.017 (3 comparisons, 0,05/3), power of 90%, and 20% loss to follow up, we would need a total of 333 participants per group.

**O�cial Title**

A Pragmatic Superiority Randomized Controlled Trial Comparing Brief Cognitive Behavioral Telepsychotherapy, Brief Interpersonal Telepsychotherapy and Telepsychoeducation for the Reduction of Emotional Distress During COVID-19 Outbreak in Professionals and Students From Essential Services With a High Level of Emotional Distress in Brazil in the Context of COVID-19

**Conditions**
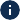

Mental Disorder

COVID

Emotional Distress

Depression

**Intervention / Treatment**
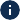

- Behavioral: Cognitive Behavioral Brief-Telepsychotherapy
- Behavioral: Brief Interpersonal Telepsychotherapy
- Behavioral: Telepsychoeducation

**Other Study ID Numbers**
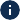

- 20200213_Treatment

**Study Start (Actual)**
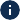

2020-11-05

**Primary Completion (Estimated)**
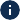

2021-07-13

**Study Completion (Estimated)**
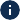

2021-07-20

**Enrollment (Estimated)**
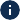

999

**Study Type**
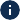

Interventional

**Phase**
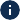

Not Applicable

**Resource links provided by the National Library of Medicine**

[MedlinePlus](https://medlineplus.gov/) (https://medlineplus.gov/) related topics: [COVID-19 (Coronavirus Disease 2019)](https://medlineplus.gov/covid19coronavirusdisease2019.html) (https://medlineplus.gov/covid19coronavirusdisease2019.html)

[Other U.S. FDA Resources](https://classic.clinicaltrials.gov/ct2/info/fdalinks) [(https://classic.clinicaltrials.gov/ct2/info/fdalinks)](https://classic.clinicaltrials.gov/ct2/info/fdalinks)

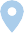

**Contacts and Locations**

This section provides the contact details for those conducting the study, and information on where this study is being conducted.

**Brazil**

**Rio Grande Do Sul Locations**

[**Porto Alegre, Rio Grande Do Sul, Brazil,**](https://clinicaltrials.gov/) [**90035-903**](https://clinicaltrials.gov/)

[Hospital de Clínicas de Porto Alegre](https://clinicaltrials.gov/)

Click to view interactive map

**Participation Criteria**

Researchers look for people who fit a certain description, called eligibility criteria. Some examples of these criteria are a person's general health condition or prior treatments.

For general information about clinical research, read [Learn About Studies](https://clinicaltrials.gov/study-basics/learn-about-studies) [(https://clinicaltrials.gov/](https://clinicaltrials.gov/study-basics/learn-about-studies) [study-basics/learn-about-studies)](https://clinicaltrials.gov/study-basics/learn-about-studies).

**Eligibility Criteria**

**Description**

Inclusion Criteria:

Professionals and students from essential services suffering from high levels of emotional distress

- T score higher than 70 on the PROMIS Anxiety Scale
- T score higher than 70 on the PROMIS Depression Scale
- T score higher than 70 on the PROMIS Anger Scale Exclusion Criteria:
- Moderate to severe suicide risk assessed by a psychiatrist

**Ages Eligible for Study**

(Child, Adult, Older Adult )

**Sexes Eligible for Study**

All

**Accepts Healthy Volunteers**

No

**Study Plan**

This section provides details of the study plan, including how the study is designed and what the study is measuring.

**How is the study designed?**

**Design Details**

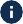

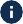

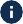

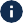

**Primary Purpose	:** Treatment

**Allocation	:** Randomized

**Interventional Model	:** Parallel Assignment

**Interventional Model Description:** Three-arm randomized clinical trial , including professionals and students from essential services suffering from high levels of anxiety, depression, and irritability symptoms during the COVID-19 outbreak randomized 1:1:1: to the Cognitive Behavioral Brief-Telepsychotherapy group (B-CBT, four sessions), Brief Interpersonal Telepsychotherapy (B-IPT, four sessions) or Telepsychoeducation group (a single session psychoeducation group plus weekly personalized pre-recorded videos for 4 weeks).

**Masking	:** None (Open Label)

**Arms and Interventions**

| **Participant Group/Arm**  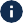 | **Intervention/Treatment** 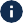 |
| --- | --- |
| Experimental: Intervention I: Cognitive Behavioral Brief- Telepsychotherapy  Four sessions of cognitive-behavioral therapy (CBT) conducted through a video call by a psychologist, accompanied by sending videos of 2 to 3 minutes with psychoeducational content and content related to CBT technique. | Behavioral: Cognitive Behavioral Brief- Telepsychotherapy   - Four sections of Cognitive Behavioral Brief- Telepsychotherapy plus personalized pre- recorded videos |
| Experimental: Intervention II: Brief Interpersonal Telepsychotherapy  Four sessions of interpersonal therapy (IPT) conducted by video call by a psychologist, accompanied by the sending of 2 to 3 | Behavioral: Brief Interpersonal Telepsychotherapy   - Four sections of Brief Interpersonal Telepsychotherapy plus personalized pre- recorded videos |

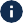

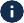

**Primary Outcome Measures**

**Secondary Outcome Measures**

**What is the study measuring?**

|  | minute videos with psychoeducational content and content related to the ITP technique. |  |
| --- | --- | --- |
|  | Active Comparator: Active Comparator: Telepsychoeducation group  One single session of psychoeducation conducted through a video call by a psychologist, accompanied by sending videos of 2 to 3 minutes with psychoeducational content for 4 weeks. | Behavioral: Telepsychoeducation   - A single session psychoeducation group plus weekly personalized pre-recorded videos for 4 weeks |

| **Outcome Measure** | **Measure Description** | **Time Frame** |
| --- | --- | --- |
| Symptom Reduction at 1- month | Proportion of participants with a 50% reduction in T-scores at 1-month in the scales Patient-Reported Outcomes Measurement Information System (PROMIS) of Depression, Anxiety and Anger that were scored above 70 at baseline. | 1-  month |

|  | **Outcome Measure** | **Measure Description** | **Time Frame** |
| --- | --- | --- | --- |
|  | Symptom | Proportion of participants with a 50% | 3 and |
|  | Reduction at 3 | reduction in T-scores at 3 and 6-months | 6- |
|  | and 6-months | follow-up in the scales Patient-Reported | month |
|  | follow-up | Outcomes Measurement Information System |  |
|  |  | (PROMIS) of Depression, Anxiety and Anger |  |
|  |  | that were scored above 70 at baseline |  |
|  |  | measured |  |
|  | Remission | Remission levels (proportion of patients with | 1, 3 |
|  | Levels in | T-score of 50 or below) in distress scales | and 6- |
|  | distress scales | (PROMIS of Depression, Anxiety and Anger) | months |
|  | Service | Service satisfaction measured by the net- | 1- |
|  | Satisfaction | promoter score at the end of the treatment | month |
|  |  | (proportion of promoters - scores of 9 or 10) |  |
|  | Improvement | Mean score change in quality of life scale | 1, 3 |
|  | in Quality of | (PROMIS General Life Satisfaction Scale or | and 6- |
|  | Life | GLSS) | months |

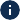

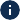

**Collaborators and Investigators**

This is where you will find people and organizations involved with this study.

**Sponsor**

**Hospital de Clinicas de Porto Alegre**

**Collaborators**

No information provided

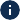

**Investigators**

- Principal Investigator: Giovanni Salum, MD, PhD, Hospital de Clínicas de Porto Alegre, Porto Alegre/Brazil

**Publications**

The person responsible for entering information about the study voluntarily provides these publications. These may be about anything related to the study.

**General Publications**

No publications available

*** Find** [**Publications about Study Results**](https://clinicaltrials.gov/) **and related** [**Pubmed Publications**](https://clinicaltrials.gov/) **in the “Results” section of the study record.**

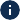

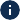

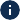

**Study Record Dates**

These dates track the progress of study record and summary results submissions to ClinicalTrials.gov. Study records and reported results are reviewed by the National Library of Medicine (NLM) to make sure they meet specific quality control standards before being posted on the public website.

**Study Registration Dates**

**Study Record Updates**

**Last Update Submitted that met**

**First Submitted**

2020-11-05

**First Submitted that Met QC Criteria**

2020-11-18

**First Posted**

2020-11-19

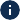

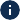

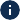

**QC Criteria**

2020-11-18

**Last Update Posted**

2020-11-19

**Last Verified**

2020-11

**More Information**

**Additional Relevant MeSH Terms**

Mental Disorders

**Terms related to this study**

**Plan for Individual Participant Data (IPD)**

**Plan to Share Individual Participant Data (IPD)?**

Yes

**IPD Plan Description**

Plan to share study protocol, SAP, ICF, CRS, analytic code and individual-based variables.

**IPD Sharing Access Criteria** Researchers and civil society **IPD Sharing Time Frame**

Avaliable six months after study completion (antecipated - January 2022)

**IPD Sharing Supporting Information Type**

Study Protocol

Statistical Analysis Plan (SAP) Informed Consent Form (ICF) Clinical Study Report (CSR) Analytic Code

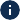

**Studies a U.S. FDA-Regulated Drug Product**

No

**Studies a U.S. FDA-Regulated Device Product**

No

**Study Documents**

No study documents available

**Drug and device information, study documents, and helpful links**
